# THE USE OF BOTULINUM TOXIN FOR SPASTICITY IN PEOPLE AFFECTED BY SPINAL CORD INJURY: A SCOPING REVIEW PROTOCOL

**DOI:** 10.1101/2024.03.30.24304908

**Authors:** Zadra Alessandro, Feller Daniel, Maini Irene, Zarbo Michele, Bonavita Jacopo

## Abstract

**Introduction:** Up to 80% of individuals affected by Spinal Cord Injury (SCI) develops spasticity, both in the acute and chronic phases. Botulinum NeuroToxin (BoNT) injections have been proven to be an effective intervention to treat focal spasticity in Stroke, Cerebral Palsy and Multiple Sclerosis. However, to the best of our knowledge, only one systematic review published in 2015 evaluated the efficacy of BoNT in treating spasticity in patients affected by SCI.

**Objective:** We formulated the following research question: “What is known from the existing literature about the use of BoNT for treating spasticity in patients affected by SCI?”.

**Material and methods:** We will include studies focusing on using BoNT to treat spasticity in patients with SCI due to any cause. We will search the following databases: MEDLINE (via PubMed), Embase, CENTRAL, Scopus, and PEDro. In addition, grey literature and the reference lists of all relevant studies will be searched. Two authors will independently perform the study selection and data extraction phases. Results from the scoping review will be summarized descriptively through tables and diagrams.

## INTRODUCTION

Up to 80% of individuals affected by Spinal Cord Injury (SCI) develops spasticity, both in the acute and chronic phases. ^1^ The definition of spasticity by Lance (1980) ^2^, largely adopted by a wide range of studies, is *“a motor disorder characterized by a velocity-dependent increase in tonic stretch reflexes (muscle tone) with exaggerated tendon jerks, resulting from hyper-excitability of the stretch reflexes, as one component of the upper motor neuron syndrome”*. However, different definitions of spasticity have been proposed since, such as Pandyan’s one ^3^, which defines spasticity as a *“disordered sensori-motor control, resulting from an upper motor neuron lesion, presenting as intermittent or sustained involuntary activation of muscles”, which may better encompass the impact of spasticity on functional movement*. Multiple mechanisms involved in the pathophysiology of spasticity are proposed, they vary depending on the lesion location, whether due to brain injury or SCI. ^4^

In recent years, the epidemiology of SCI has shown profound changes leading to an increase in lesions due to non-traumatic causes, incomplete and with cervical Neurological Level of Injury (NLI). ^5^ This epidemiologically evolving scenario poses new challenges for rehabilitation practitioners: spasticity often presents as a focal pattern in incomplete lesions and may be treated with focal interventions. Generally, limb spasticity is managed through physical interventions (stretching and bracing), oral or intrathecal medications (Baclofen, Tizanidine) and surgery. ^6^ Oral drugs are unfortunately limited in their use due to side effects like sedation, dizziness, confusion, generalized weakness and hypotension. ^7^ Intrathecal administration is usually indicated for diffuse spasticity. ^8^ For these reasons, chemodenervation via local intramuscular injections of Botulinum NeuroToxin (BoNT) may be used to treat focal spasticity while minimizing systemic side effects. ^9^

Spasticity hinders function in activities of daily living, including self-hygiene, can be a source of pain and fatigue and plays a role in the development of contractures and pressure sores. Spasticity can also negatively affect quality of life and participation. ^10^ However, some literature ^6^ suggests that spasticity in SCI could also be beneficial in increasing stability in sitting and standing, improving muscle bulk, and preventing deep vein thrombosis, osteopenia and osteoporosis. These findings, therefore, suggest that it is critical how spasticity assessment is performed. Recent reviews ^11^ reported that subjects with SCI do not always receive adequate treatment because of an inadequate assessment. Clinical evaluation of spasticity should consider patient preferences and include patient-reported measures to capture the impact on activities of daily living and quality of life, enabling shared treatment goals between patient and clinician.

BoNT injections have been proven to be an effective intervention to treat focal spasticity in Stroke, Cerebral Palsy and Multiple Sclerosis. ^12–14^ In SCI, BoNT has found a consolidated indication in neurogenic detrusor overactivity treatment ^15^, while its use for spasticity is currently an evolving field of research and clinical practice. To the best of our knowledge, the most recent systematic review ^16^ aimed to assess the efficacy of BoNT chemodenervation, phenol and alcohol neurolysis in people affected by spasticity following SCI dates back to 2015 and has shown the lack of high-quality evidence, advocating for further research in the field. In the same review, the authors didn’t find any randomized or non-randomized controlled trials; therefore, we opted to map the existing literature with a scoping review design.

## OBJECTIVES

We formulated the following research question: “What is known from the existing literature about using BoNT for treating spasticity in patients following SCI?”.

In particular, we will aim to determine from every included study:

I. The adopted definition(s) to diagnose spasticity;
II. The goals of spasticity treatment;
III. The outcome measures employed to assess patients with spasticity and SCI;
IV. The use of BoNT to treat spasticity regarding target muscles, dosage, botulinum toxin type, guidance for injection procedures;
V. The efficacy of treatment with BoNT, compared to other interventions or placebo;
VI. The study design employed.

## MATERIALS AND METHODS

The present scoping review will be conducted in accordance with the JBI methodology for scoping reviews. ^17^ The Preferred Reporting Items for Systematic Reviews and Meta-Analyses extension for Scoping Reviews (PRISMA-ScR) Checklist for reporting will be used. ^18^

### Inclusion criteria

Studies will be eligible for inclusion if they meet the following Population, Concept, and Context (PCC) criteria:

- Population. We will include patients 18 years old or older with SCI due to any cause affected by spasticity not attributable to traumatic brain injury, cerebral palsy, stroke, multiple sclerosis, degenerative disorders or hereditary pathologies.
- Concept. Studies should focus on BoNT as an intervention administered intramuscularly with any guidance, such as electromyography or ultrasound. We will include experimental studies independently from the comparator used.
- Context. This review will consider studies conducted in any context.

### Sources

This scoping review will consider any study design or publication type written in English. We will only include secondary studies (e.g., systematic reviews) with a foreground question. No time, geographical or setting limitation will be applied. Studies could be quantitative, qualitative or mixed-method in order to consider different aspects of spasticity burden across several domains. We will include studies focusing on a mixed population when the majority of the population (> 50%) are patients with SCI. We will also include studies presenting a separate SCI group analysis.

### Search strategy

We will search the following databases: MEDLINE (via PubMed), Embase, CENTRAL, Scopus, and PEDro. MEDLINE (via PubMed) will be searched using the following search string:

*(((spinal OR spine) AND (cord OR injur* OR lesion* OR trauma* OR transaction* OR laceration* OR contusion*)) OR myelopath* OR SCI OR paraplegia OR paraparesis OR quadriplegia OR quadriparesis OR tetraplegia OR tetraparesis) AND (spasticity OR spastic OR hypertonia OR clonus OR co-contraction OR spasm* OR “associated reaction*” OR withdrawal) AND (botulinum OR bont* OR botox OR xeomin OR dysport OR onabotulinum OR incobotulinum OR abobotulinum)*

In addition, grey literature (i.e., Google Scholar), trial registers (i.e., ClinicalTrials.gov and the WHO International Clinical Trials Registry Platform portal) and the reference lists of all relevant studies will be searched. Finally, we will contact experts in the SCI field (top ten experts on SCI according to ExpertScape.com on 11/03/2024) asking for additional references not retrieved in the database search.

### Study selection

Duplicates will be removed using the deduplication function of “Systematic Review Accelerator.” ^19^ Two researchers will independently perform the study selection process, firstly by title/abstract and finally by full text. Any disagreement will be solved by consensus or by the decision of a third author. We will use the online electronic systematic review software package (Rayyan QCRI) to organize and track the selection process. ^20^

### Data extraction

Data will be extracted using two Word templates, one for primary and one for secondary studies. The data extraction form will be piloted in three primary and secondary studies.

The data extraction process will be conducted independently by two reviewers. Any discrepancies will be resolved by a consensus between the two authors and eventually by a third author’s decision.

### Data synthesis

Results from the scoping review will be summarized descriptively through tables and diagrams. Specifically, we will summarize and synthesize the data extracted from the studies and highlight any knowledge and implementation gaps for using BoNT in patients with SCI.

## Data Availability

All data produced in the present work will be contained in the final manuscript

